# Age-specific social mixing of school-aged children in a US setting using proximity detecting sensors and contact surveys

**DOI:** 10.1101/2020.07.12.20151696

**Authors:** Kyra H. Grantz, Derek A.T. Cummings, Shanta Zimmer, Charles Vukotich, David Galloway, Mary Lou Schweizer, Hasan Guclu, Jennifer Cousins, Carrie Lingle, Gabby M.H. Yearwood, Kan Li, Patti Calderone, Eva Noble, Hongjiang Gao, Jeanette Rainey, Amra Uzicanin, Jonathan M. Read

## Abstract

Comparisons of the utility and accuracy of methods for measuring social interactions relevant to disease transmission are rare. To increase the evidence base supporting specific methods to measure social interaction, we compared data from self-reported contact surveys and wearable proximity sensors from a cohort of schoolchildren in the Pittsburgh metropolitan area. Although the number and type of contacts recorded by each participant differed between the two methods, we found good correspondence between the two methods in aggregate measures of age-specific interactions. Fewer, but longer, contacts were reported in surveys, relative to the generally short proximal interactions captured by wearable sensors. When adjusted for expectations of proportionate mixing, though, the two methods produced highly similar, assortative age-mixing matrices. These aggregate mixing matrices, when used in simulation, resulted in similar estimates of risk of infection by age. While proximity sensors and survey methods may not be interchangeable for capturing individual contacts, they can generate highly correlated data on age-specific mixing patterns relevant to the dynamics of respiratory virus transmission.

## Introduction

Social interactions or person-to-person contacts can influence the transmission of many infectious diseases. Respiratory viruses, such as influenza, are transmitted primarily through large infectious droplets when an ill person coughs or sneezes within relatively short distances of others (1, 2). Close contact facilitates transmission of respiratory diseases, but the extent to which various populations are connected by these potential transmission events remains unclear. Identifying more precisely the routes of disease transmission therefore has important implications for public health policy and pandemic response, and can direct resources to most efficiently target groups at high risk of transmission.

The POLYMOD study (3) was the first large-scale, survey-based study aimed at quantifying social contact patterns in eight European countries, showing differential contact rates by age and age-assortative mixing. The empirical data on social interactions and contact behaviours collected through POLYMOD and other survey-based studies have improved efforts to explain and predict spread of infectious disease, including mumps and influenza (4–10). Many studies since have sought to characterize social mixing patterns in a variety of populations using contact surveys (11–28) as well as wearable proximity sensors (11, 22, 23, 29–33), social media or mobile phone data (22, 34, 35), direct observation (36), and model-based approaches incorporating demographic and time-use data (37–39). Few studies, though, have considered the use of non-survey social contact data in parameterizing mathematical models (9, 40).

There has been particular interest in the role of school-aged children (approximately 5–18 years of age) in transmission of many respiratory infections (41). Schoolchildren are at high risk of infection by influenza and other respiratory pathogens (42, 43). The local nature of geographic spread during the 2009 A/H1N1 pandemic, the strong associations between pandemic onset and school openings, and the high attack rates observed within schools all confirm the critical role schoolchildren play in facilitating transmission (44–47). Schoolchildren generally display highly assortative mixing by age (i.e., they preferentially interact with children of the same age) and high contact rates with adults and the elderly (their parents and grandparents) which may facilitate transmission among schoolchildren and within their surrounding communities (3, 4, 13, 16, 25, 48). Many public health interventions, including school closures and vaccination campaigns, focus on the role of schoolchildren in the spread of respiratory infections (49, 50).

One challenge in drawing links between patterns of social contacts and respiratory disease transmission is the difficulty in empirically measuring patterns of proximal social interaction. Social contacts that can lead to transmission of pathogens can potentially be transient, non-synchronous (i.e., through contamination of the environment), and of varying intensity (2, 51). Multiple methods have been used to measure social contact, the relative disadvantages and advantages of which have been described elsewhere (51). The majority have used interviews or surveys to collect data on self-reported contacts, raising the possibility of significant recall bias (52). These contacts may or may not involve conversation, physical touch, or other features that may make them more memorable and thus more likely to be reported. Proximity-detecting wearable sensors, or motes, offer an alternative to self-reported data by automatically logging other sensors when nearby (31). These proximal interactions, though, may differ in important ways from self-reported interactions and may not capture aspects of social interactions that are critical to the transmission of respiratory pathogens. Little information is available on the comparative reliability and limitations of these methods in characterizing epidemiologically important social contacts (11, 52–56).

To support the use of social contact data to inform epidemic models, we conducted paper and online contact surveys and deployed proximity sensors in a population of US schoolchildren. We compared individual-level and aggregate age-specific mixing patterns captured by both methods and considered qualitative and quantitative differences in predicted attack rates from transmission models of respiratory pathogens using these data.

## Results

### Study population and average contact patterns

The Social Mixing and Respiratory Transmission (SMART) study was conducted in eight schools in the Pittsburgh standard metropolitan statistical area from February 2012 to June 2012. Students in kindergarten (K) to 12^th^ grade were eligible to participate. At least one contact survey distribution overlapped with a proximity sensor deployment in each school, and students could participate in more than one contact survey distribution.

Of the 2,337 students enrolled in the eight participating schools, 1,325 (56.7%) completed 2,155 contact surveys, and 1,834 (78.5%) participated in a school-based sensor deployment (Table 1). Of the students who participated in a sensor deployment, 1,125 (61.3%) completed at least one contact survey, 826 (45.0%) students completed a survey about a day on which they also wore a sensor, and 730 (39.8%) completed two surveys. Generally, younger students were overrepresented in the sensor deployment populations and underrepresented in the survey-completing populations (Fig. 1, Supp. Table 1).

**Figure 1.**
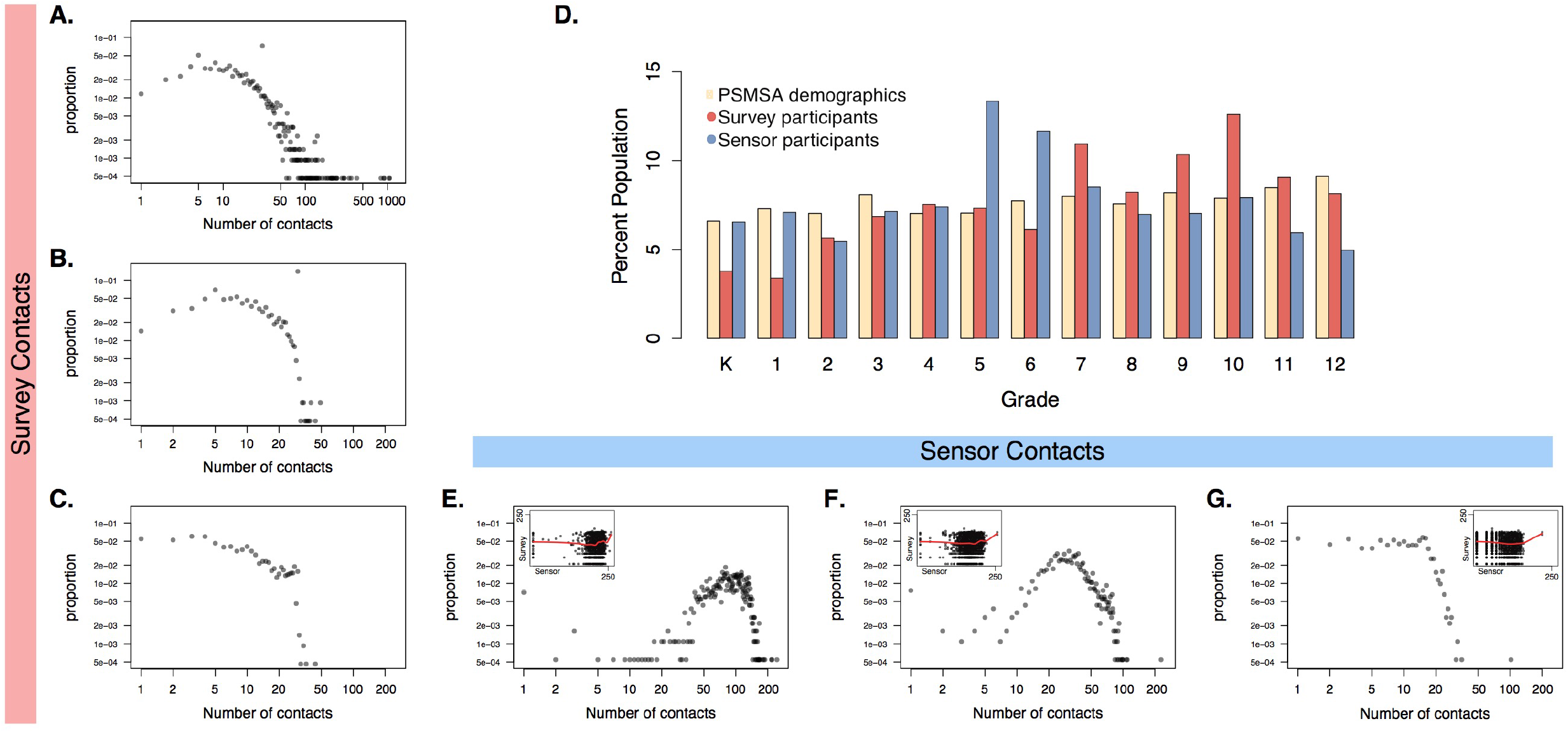
Distribution of the number of contact events recorded in a US school setting by self-reported contact surveys and proximity-detecting sensors: (A) total survey-reported contacts; (B) detailed survey-reported contacts; (C) survey-reported in-school contacts; (E) all unique contacts recorded by sensors; (F) all unique contacts with more than 10 cumulative contacts (roughly 3 minutes of interaction); and (G) all unique contacts with more than 100 cumulative contacts (roughly 30 minutes of interaction). Insets in (E)-(G) show the plot of in-school survey contacts versus each metric of sensor-recorded contacts with a cubic smoothing spline. (D) shows the population distribution by grade of participants who completed at least one contact survey or participated in a sensor deployment, compared to the population distribution of the Pittsburgh standard metropolitan statistical area (PSMSA) for 2012.

**Table 1.** Study population and average number of contacts recorded by self-reported contact surveys and proximity-detecting sensors in a US school setting

| Grade | K | 1 | 2 | 3 | 4 | 5 | 6 | 7 | 8 | 9 | 10 | 11 | 12 | Total |
| --- | --- | --- | --- | --- | --- | --- | --- | --- | --- | --- | --- | --- | --- | --- |
| Completed ≥ 1 contact survey |  |  |  |  |  |  |  |  |  |  |  |  |  |  |
| No. participants | 49 | 46 | 75 | 91 | 100 | 97 | 81 | 146 | 108 | 134 | 171 | 119 | 108 | 1325 |
| No. survey responses | 53 | 55 | 110 | 175 | 143 | 175 | 139 | 219 | 176 | 197 | 302 | 212 | 199 | 2155 |
| Mean no. detailed contacts (sd) | 5.0<br>(2.2) | 3.7<br>(2.3) | 9.7<br>(6.5) | 11.0<br>(7.2) | 15.2<br>(8.0) | 10.8<br>(8.2) | 16.5<br>(9.5) | 18.7<br>(9.7) | 16.0<br>(9.4) | 17.2<br>(9.3) | 13.9<br>(9.0) | 14.3<br>(9.0) | 16.0<br>(10.7) | 14.2<br>(9.4) |
| Participated in a sensor deployment |  |  |  |  |  |  |  |  |  |  |  |  |  |  |
| No. participants | 119 | 131 | 100 | 131 | 136 | 245 | 215 | 157 | 127 | 125 | 149 | 108 | 91 | 1834 |
| Mean no. unique contacts (sd) | 84.6<br>(31.4) | 81.2<br>(22.6) | 81.8<br>(29.0) | 81.3<br>(28.0) | 83.1<br>(24.5) | 83.2<br>(25.6) | 91.8<br>(28.4) | 92.8<br>(25.8) | 76.3<br>(33.7) | 109.0<br>(32.9) | 109.6<br>(35.1) | 111.9<br>(38.2) | 102.5<br>(36.0) | 90.8<br>(31.9) |
| Completed ≥ 1 contact survey and participated in sensor deployment |  |  |  |  |  |  |  |  |  |  |  |  |  |  |
| No. participants | 43 | 43 | 65 | 84 | 88 | 81 | 55 | 129 | 82 | 119 | 143 | 106 | 87 | 1125 |
| No. survey responses | 46 | 51 | 99 | 163 | 131 | 149 | 104 | 192 | 134 | 179 | 258 | 190 | 159 | 1855 |
| No. participants with sensor-day survey | 0 | 0 | 28 | 66 | 65 | 68 | 53 | 56 | 61 | 118 | 129 | 99 | 83 | 826 |

Detailed survey-reported contacts are those interactions for which a student also reported contact age, sex, duration, and context. Unique sensor-recorded contacts for each participant is the total number of other participants with whom their proximity sensor recorded at least one interaction during a sensor deployment. Sd, standard deviation.

Proximity sensors on average captured more contacts, defined as the total number of participants with whom an individual recorded at least one sensor interaction, per student than self-reported surveys, which captured self-reported social contacts with all students, including interactions which involved speaking, playing, or touching (Table 1, Supp. Table 1). The range in the number of survey-reported contacts, particularly for total contacts, was large (Fig. 1). Few paper surveys reported more than 30 contacts per day (0.5%, 8/1760), compared to web-completed surveys (3.0%, 12/395). The distribution of the number of unique sensor-recorded contacts was less skewed, but the presence of several high-degree nodes (individuals with many contacts) became increasingly apparent as the minimum number of cumulative contacts (an approximation of contact duration) required to be considered a unique contact was increased. The average duration of a survey-reported contact was 124.3 minutes, compared to just 7.5 minutes for sensor-recorded contacts. There was marked similarity between the distribution of survey-reported in-school contacts (Fig. 1C) and unique sensor-recorded contact events with at least 100 cumulative contacts (Fig. 1G), but the association at an individual level was unclear.

### Individual-level concordance of contact surveys and sensors

In multivariate regression analysis adjusted for participant age, sex, and survey design, sensor-recorded and survey-reported contacts rarely served as significant predictors of one another (Fig. 2, Supp. Fig. 1). Increasing the cumulative contact threshold for sensor contacts did not improve these associations. Generally, the number of survey-reported contacts increased with age. Duration of survey-reported contacts increased with age as well, but the effect size was reduced compared to the number of contacts. Survey type or method of administration was not associated with number or duration of recorded contacts. Male students were less likely than female students to report contacts and reported shorter contacts on average in contact surveys.

**Figure 2.**
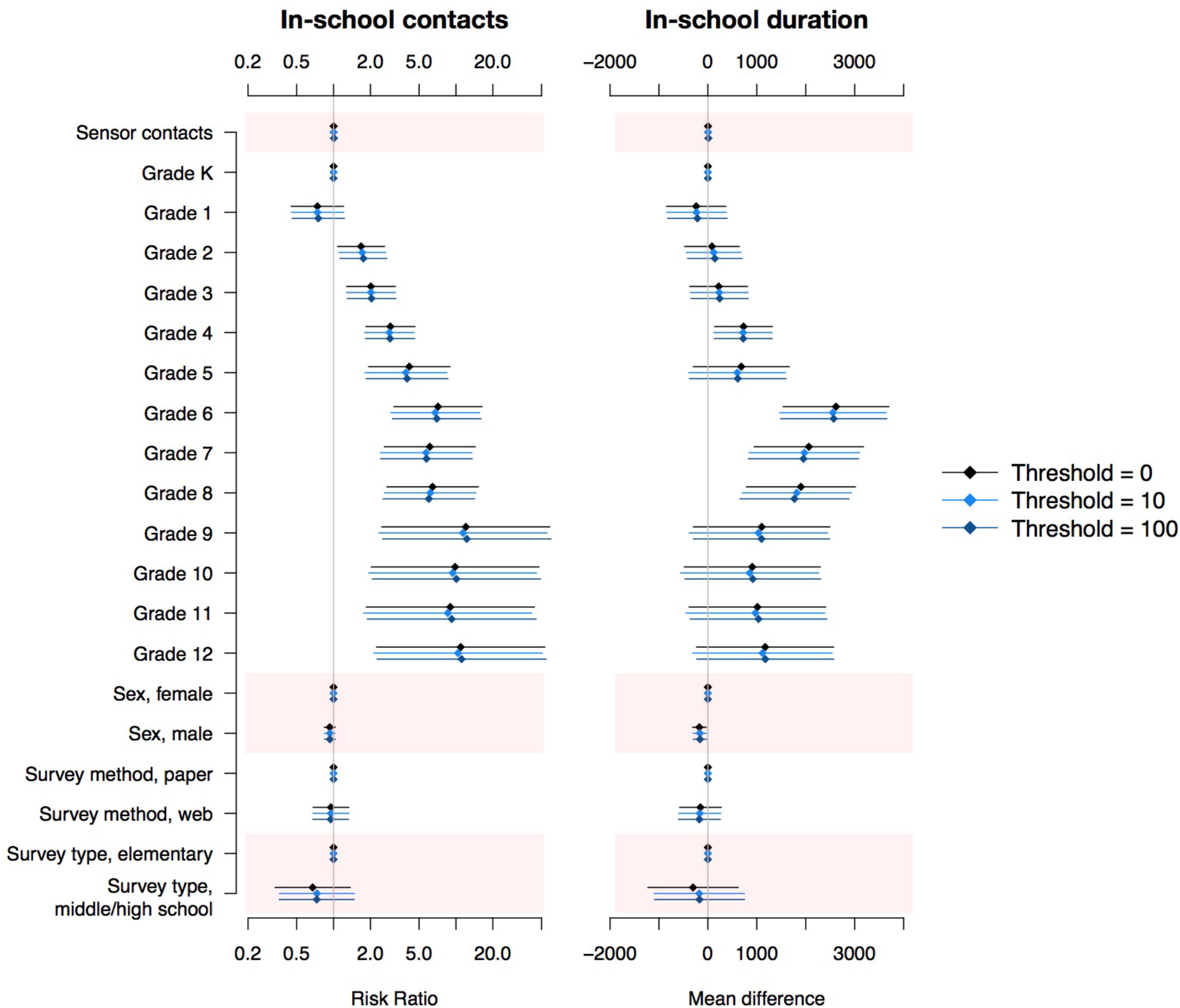
Factors associated with the number and duration of survey-reported in-school contacts in a US school setting. All models include a random intercept for day of survey completion.

Results using multiple thresholds of cumulative sensor contact are shown in the supplement (Supp. Fig. 2). We found significant associations between sensor outcomes and number of survey-recorded contacts; however, the effect size was small relative to other factors (e.g., age).

### Age-specific mixing patterns

Age-specific contact patterns derived from both data collection methods showed highly assortative mixing. For example, in contact surveys, participants reported up to eight times as many in-school contacts with students of the same grade than would be expected under proportionate mixing assumptions (Fig. 3A). There was also a striking consistency between pairwise survey- and sensor-recorded contact ratios as a function of the difference in grade. The average departure from proportionate mixing expectations for participants in the same grade was 4.07, compared to just 0.72 for participants one grade apart and 0.15 for participants two or more grades apart (Fig. 3E).

**Figure 3.**
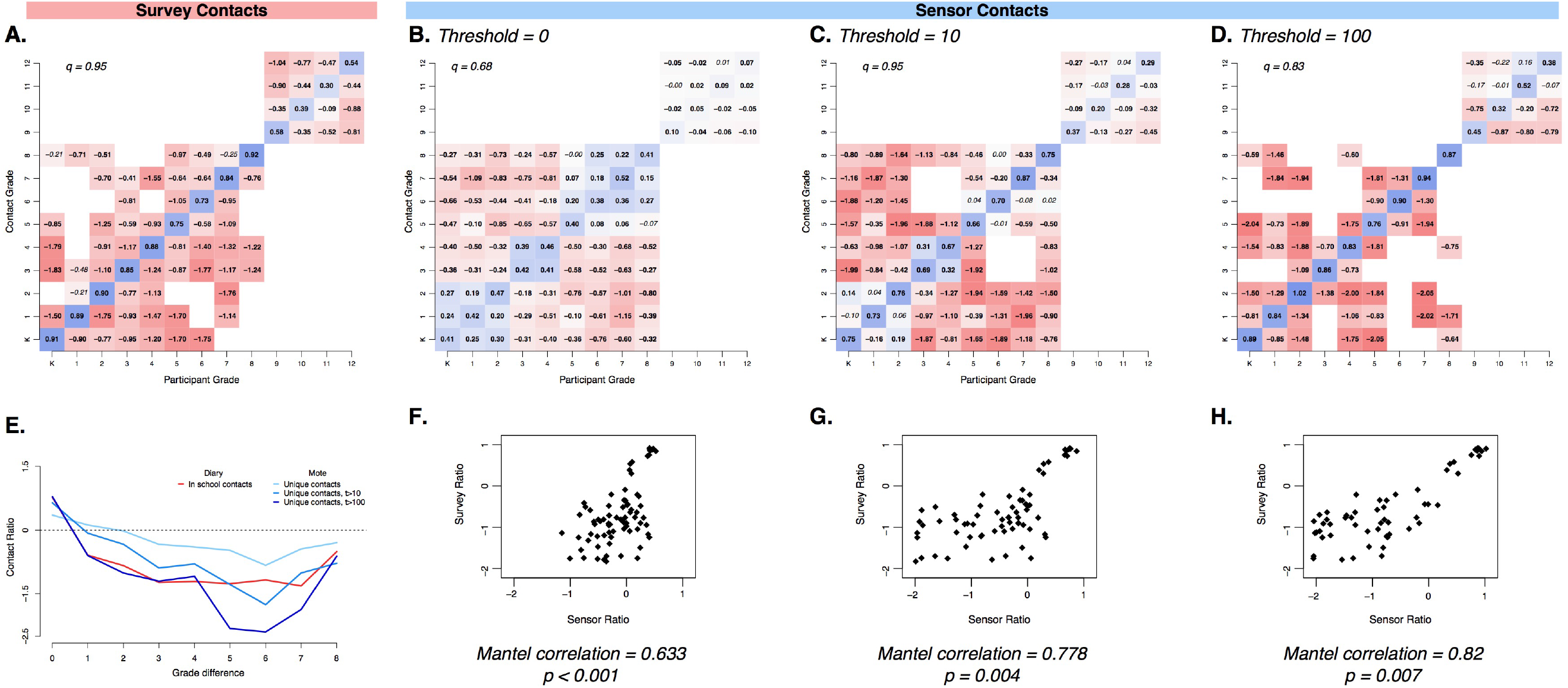
Age-specific mixing matrices generated from in-school survey contacts and unique sensor-recorded contacts in a US school setting at various cumulative contact thresholds. Matrices are presented as log-10 ratio of observed contacts relative to expectation under proportionate mixing assumptions for survey-reported in-school contacts (A) and sensor-recorded unique contacts with thresholds of 0 (B), 10 (C), and 100 (D) cumulative contacts. Blue colours indicate more contacts than expected under proportionate mixing assumptions, and red colours indicate less mixing than expected. Bolded ratio values deviate significantly from the null expectation, a=0.05, and *q* equals the degree of assortative mixing. Scatterplots (F-H) show the corresponding *i,j* values of the survey- and sensor-based mixing matrices at each threshold (0, 10, 100). (E) shows the average departure from proportionate mixing as a function of difference between grade for each matrix.

Assortativity of age-specific matrices based on contact surveys and sensor data ranged from *q* = 0.68 to *q* = 0.95 (Fig. 3). The range was partially due to the structure of the participating schools; in this study, there were no schools with both high school and non-high school students. However, even within each school, mixing patterns showed high degrees of assortative mixing (e.g., in-school contact survey-based matrices range from *q* = 0.62 to *q* = 0.99, Supp. Fig. 2).

The effect of school structure on mixing patterns was most apparent in matrices based on unique sensor contacts, which revealed three elementary grade clusters (K–2, 3–4, 5–8) within which there was strong assortative mixing (Fig. 3B). High school students (grades 9 to 12) represented a well-mixed, modular cluster (*q* = 0.05 and 0.12 for HS1 and HS2, Supp. Fig. 3). Assortativity increased when a threshold of cumulative sensor contacts was applied (Fig. 3C, 3D), as did the correlation between the age-specific contact ratios of survey- and sensor-based matrices (Mantel correlation coefficients 0.63 to 0.88). Matrices based on rate of contact (that is, unadjusted for proportionate mixing expectations) were qualitatively similar to ratio-based matrices but, as they did not account for school demography and participation rates across grades, were poor correlates between the two methods. Age-specific matrices based on survey-recorded contact events lasting longer than 10 minutes, contact events reported on days of sensor deployments, and survey- and sensor-recorded contact durations displayed similar patterns of age assortativity (Supp. Fig. 4, 5).

### Transmission models

When used in age-specific simulation, sensor- and survey-based mixing matrices produced similar attack rates when adjusted by proportionate mixing expectations (Fig. 4). Increasing the contact threshold resulted in more heterogeneity relative to the proportionate mixing baseline. There was discordance between the sensor- and survey-based predicted attack rates in particular schools, which increased with cumulative sensor contact threshold and disjuncture in contact matrices. However, in other schools, there was a marked degree of similarity between attack rates regardless of contact matrix employed. In simulations based on unadjusted contact rate matrices, predicted attack rates were lower in younger children when using survey-based matrices, a reflection of the different reporting rates by age and specific demography of each school (Supp. Fig. 6). We explored multiple parameters in our transmission model, assuming reproductive numbers of 1.5, 2 and 3. We found little qualitative difference between these simulations (Supp. Fig. 7).

**Figure 4.**
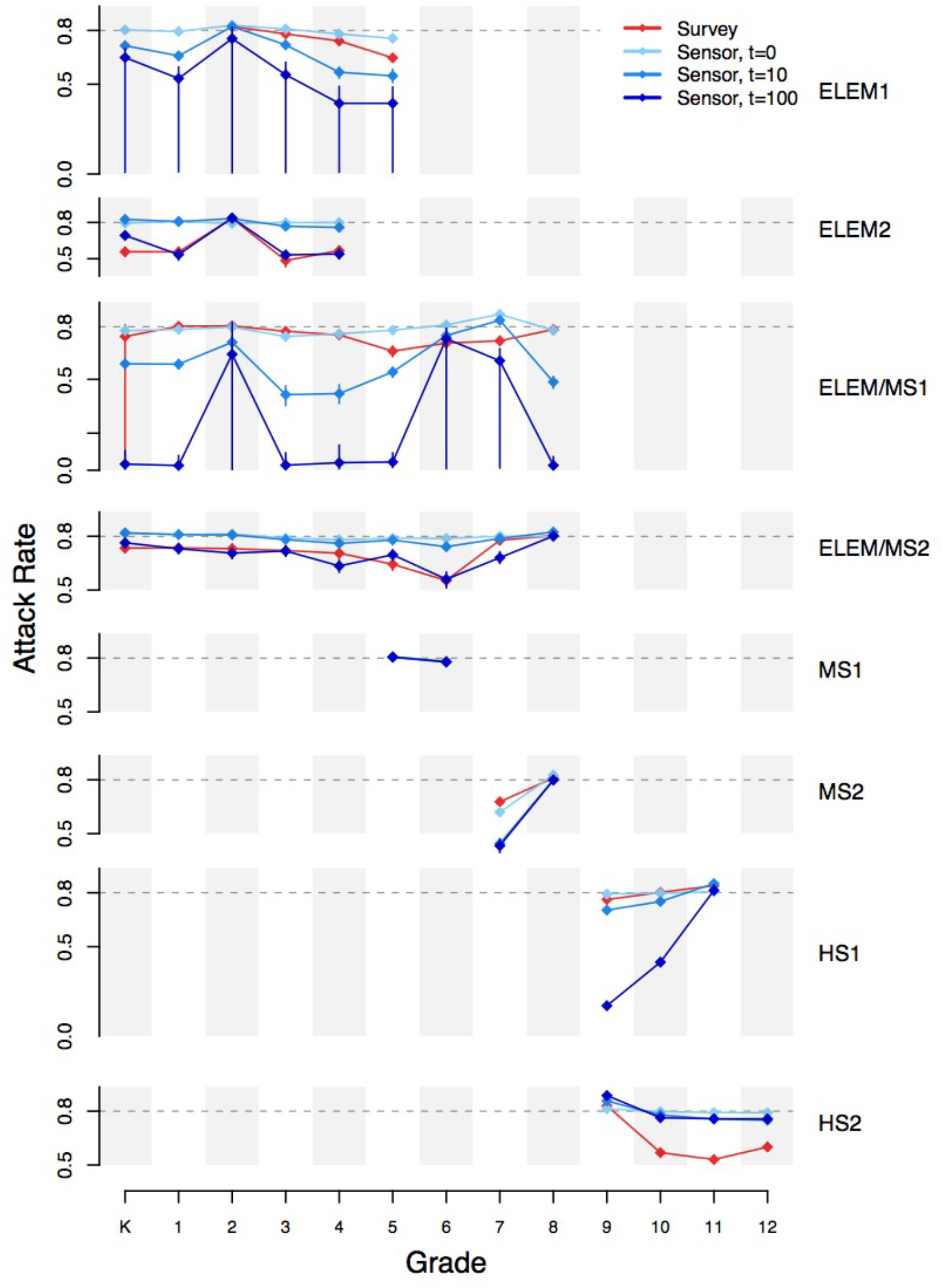
Grade-specific final predicted attack rates of a respiratory virus in a US school setting, based on stochastic simulation using mixing matrices of in-school survey contacts and unique sensor-recorded contacts at various contact thresholds, adjusted by proportionate mixing expectations, within each school (ELEM, elementary; MS, middle school; HS, high school).

## Discussion

The utility of social contact data to the study of infectious diseases has been limited in part by questions of how to best measure social interactions relevant to transmission. In this project, we found that, while the two commonly used methods captured different information at the individual level, they gave similar results in several aggregate patterns of contact that are thought to be relevant to pathogen transmission, namely, patterns of age-specific mixing and probability distributions of the total number of contacts. As in other work, we found evidence for strong assortativity of contacts by grade (32). This work has important implications for the empirical parameterization of mathematical models of transmission, particularly of respiratory pathogens. This work suggests that either empirical approach could be used to characterize age-specific interactions suitable for use in modelling to inform policy.

Previous studies (11, 22, 23, 57) which compared contact surveys to proximity sensors also found poor individual-level concordance between the two methods: anywhere from 15% to 96% of contacts captured through proximity sensors were not recorded in contact surveys. Despite the poor individual-level comparisons in these studies, some found agreement in aggregate patterns across age ranges using two different methods (30, 57).

We also observed substantial absolute differences in the number and type of contacts recorded by self-reported contact surveys and proximity sensors. We found either metric was a poor predictor of the other, even when adjusting for age, sex, and study factors. However, we found stronger individual-level correspondence between the measures when we restricted sensor data to contacts with longer cumulative duration (true for 3-minute and 30-minute minimum thresholds), consistent with earlier work which found longer contacts were more likely to be reported in surveys (11, 22, 23). In practice, the two methods are designed to capture different social interactions. Per the study protocol, survey-recorded contacts should only have included those with interactions that involved talking, playing, or touching, while sensors recorded all other sensors within proximity regardless of whether participants were socially interacting. That the correspondence increased when limiting sensor information to proximal contacts with longer duration suggests that these were more likely to be contacts which include social interactions. It is unclear which type of contact (proximal or social interaction) is most relevant for the spread of respiratory pathogens.

To determine whether contact patterns measured using different empirical approaches lead to different transmission dynamics, we simulated transmission using models parameterized with data from the two empirical techniques. In simulations using mixing matrices adjusted by proportionate mixing expectations, similar age-specific infection patterns were found using sensor and survey data. Previous work has similarly found that, while simulations using unadjusted contact data from surveys and proximity sensors differ, appropriate adjustment to survey data which capture key structural elements of the contact network (e.g., age assortativity) leads to consistent simulation results using both kinds of contact data (57). Here, differences in attack rates appear to be driven by increasing disjuncture between grades and age assortativity in certain mixing matrices.

Importantly, the metric we used to compare age-specific contact patterns from survey- and sensor-recorded data did not account for absolute differences in the overall contact rates of children in each grade. In simulation, the β estimation procedure (see *Supplementary Methods)* scaled the overall rate of contact between age-specific contact matrices, but did not account for possible age-specific differences in average contact rates. Absolute difference in contact probability may have important epidemiological consequences, particularly when considering onward transmission to the community. Transmission to family and community members outside school is a critical component of schoolchildren’s key role in respiratory disease transmission which we do not consider here. Furthermore, contact patterns are likely to be different on school holidays, weekends, or even when a child has symptomatic illness (13, 58, 59); the feasibility of electronic sensors in these contexts has yet to be shown. Future studies linking the mixing patterns and incidence of respiratory disease among schoolchildren with disease risk in their communities would provide valuable evidence for planning and control measures.

Our study has some important limitations. Though we adjusted for the demographics of the specific schools and deployments that we conducted, our results may not be generalizable to other settings. The physical and architectural environment of our schools, the density of sensors that we were able to deploy in our schools, and the specific days that we deployed our study may all have affected our results. Technical issues, though not common, did occur with the sensors, resulting in lost data for some sensors. Similarly, recall bias and misclassification by participants when completing contact surveys may have obscured the relationship between our two methodological measurements. We found that the design and administration of contact surveys led to some censoring in the number of contacts reported (Fig. 1). Nonetheless, we believe that the relationships we found were robust to the misclassifications and biases that may be generated by these sources.

Previous work has indicated that risk of infection with influenza is more closely linked to the average mixing patterns of an individual’s age group, rather than the individual’s contact behaviour (7). We found that two common methods of collecting social contact data, self-reported surveys and proximity sensors, recorded qualitatively and quantitatively different individual social mixing behaviour but could still generate similar aggregate age-specific social contact patterns. The collection of high-quality social contact data through either method has important implications for surveillance, prediction, and prevention of respiratory virus transmission. Our finding that these two methods found some commonality in aggregate age-specific social contact patterns suggests that these phenomena are not an artefact of either specific empirical method but attributes of these study populations.

## Methods

### Study description

Enrolment in the Social Mixing and Respiratory Transmission (SMART) study operated on an opt-out basis, and all students registered in a participating school before the start of the study were eligible to participate. Students in kindergarten (typically aged 5 years) to 12^th^ grade (typically aged 18 years) from two elementary (K to 4^th^ grade, K to 5^th^ grade), two middle (5^th^ to 6^th^ grade, 7^th^ to 8^th^ grade), two elementary-middle (K to 8^th^ grade), and two high (both 9^th^ to 12^th^ grade) schools were eligible to participate in SMART. Participation rates were high in all schools (82 to 99%). Each school provided aggregate demographic information about the school population, and individual grade and sex of participating students.

### Proximity sensor deployments

The details of proximity sensor deployments have been described in detail elsewhere (60). In brief, participating students were given proximity sensors in plastic pouches and instructed to wear the pouch around their neck for the duration of the school day without removing or otherwise tampering with the sensor. In six of the eight schools, all participating students were given a sensor; in two schools, the large student population limited the deployment to randomly selected classrooms in each grade. Deployments typically lasted from the first class period (08:00 – 09:00) to the last class period (14:00 – 15:00). Deployment days in each school were chosen to be representative of a typical school day, without any special schoolwide or grade-specific activities that could modify normal contact patterns.

We used TelosB wireless sensors (61) programmed in the NesC language to send beacons every 20 seconds (beacon frequency 3 per minute). The receiving sensor recorded the contacting sensor’s identity, an internal time stamp, and a radio strength signal indicator (RSSI). Signal strength provided an estimate of physical proximity, but was highly dependent on the orientation of the two sensors and any obstructions between them and therefore could not be used to define an exact distance between contacts. Based on pilot studies and previous work on effective distances of respiratory virus transmission (29, 62), we chose a signal threshold (−80 dBm) that should correspond to contacts of relevance to respiratory disease transmission.

The number of unique proximity sensor contacts recorded for a participant was defined as the total number of other participants with whom their proximity sensor recorded at least one interaction during each deployment. To explore patterns of contacts of varying length, we considered several values of the contact threshold, or the minimum number of recorded interactions between two proximity sensors required to be considered a unique contact. The number of interactions between any given pair of sensors was taken to be the maximum number of interactions recorded by either sensor, to account for battery failure, measurement error, or other malfunctions.

### Contact survey design

Contact surveys were completed by participants in school under the supervision of project staff and teachers. Each sheet of the paper version allowed for information on up to 30 contacts to be recorded; additional sheets could be requested. Two versions were designed: one for middle- and high-school students, and a simplified version for elementary school students (although some elementary school children completed the middle- and high-school version, upon consultation with school administrators and teachers). Classrooms were randomly selected to participate from each grade, and students of several classrooms completed more than one contact survey over the course of the study period.

Participants were asked to report information about any individual they talked with, played with, or touched the previous day, including the contact’s age and sex, whether they attended the same school as the participant, the context in which the contact was made, whether the contact involved direct or indirect (through a shared object) touch, and approximate duration of the contact. Students reported the total number of contacts made in the previous day, without detailed information, and additional demographic information about themselves and their household. The surveys were completed either on paper or by computer, depending on resources available in each school.

We defined total survey contacts as the total number of individuals a student reported having interacted with on the day before the survey was completed. Detailed contacts were the subset of total contacts for which the student reported contact age, sex, duration, and context. We considered further subsets of detailed survey contacts, including those occurring within school, those reported to have lasted more than 10 minutes over the course of the day, and those occurring on the same day as a sensor deployment.

### Statistical analysis of individual contact patterns

Transmission risk is likely dependent on both the type and length of interaction. We therefore estimated contact durations for survey- and sensor-recorded contacts using an exponential fitting method, following the work of Read and Danon (12, 18) (*Supplementary Methods*). Briefly, each sensor interaction was assumed to represent an independent contact of between 0 to 20 seconds; the total interactions between a pair of participants were summed to compute the total duration of contact in one deployment. Participants were asked to record the approximate durations of survey-reported contacts.

We used negative binomial regression to investigate which factors were associated with the number of reported contacts for each student who participated in a sensor deployment and completed at least one contact survey. Each model included participant grade, gender, and a random intercept term for day of survey completion or sensor deployment. Survey administration and sensor deployment days were unique to each school. Terms for the type and method of survey administration were added to models of survey-recorded outcomes.

### Age-specific mixing matrices

We estimated two metrics of age-specific contact patterns: an average per-capita mixing rate, and the age-specific mixing ratio of observed contact rates to those expected under the assumption of proportionate mixing.

### Average per-capita mixing rate

The first metric is the average number or duration of contacts recorded or reported by a participant in any grade *i* with a student in grade *j*:

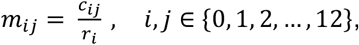

where c_i,j_ is the total number or duration of contacts recorded by participants of grade *i* with individuals in grade *j*, divided by the number of participants in grade *i*, r_i_. In sensor deployments, contacts could only be recorded with other participants (sensor-wearers) in grade *j*. If a certain grade was underrepresented among participants relative to other grades, fewer contacts would be recorded with individuals in that grade because of the low sensor coverage. Therefore, the sensor contact rate was adjusted by the ratio of the proportion of the total school population in grade *j* to the proportion of sensor participants in grade *j*, 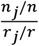, where *n*_*j*_ is the number of students in grade *j* regardless of participation.

### Age-specific mixing ratio

We also estimated the ratio of the observed age-specific contact rate to the expected contact rate if the probability of contact were dependent solely on the availability of potential contacts in a given grade (proportionate mixing assumption).

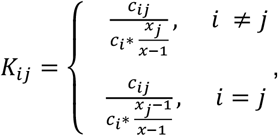

where x_j_ is the number of individuals in grade *j* with whom participants in grade *i* could record a contact, and all other terms are as defined above. Values greater than 1 indicate more contacts were recorded by participants in grade *i* with individuals of grade *j* than would be expected under proportionate mixing. Proportionate mixing assumes that an individual in grade *i* mixing at random will contact individuals in grade *j* with a probability equal to the proportion of the population in grade *j*, but no assumption is made on the probability of individuals in grade *i* making any contact relative to other groups.

By design, *r*_*j*_, the participant population, is equal to *x*_*j*_, the contact population, in sensor deployments. For within-school contacts, we used the demographic information of all registered students in each school to define the potential contact population. Combined K-12 matrices were generated by averaging age-specific matrices from all participating schools, weighted by the number of participants in each school.

Confidence intervals were calculated using 1,000 resampled bootstrap replicates of contact events. Mantel correlation coefficients were used to compare mixing matrices. The degree of assortative mixing, *q*, was calculated as the ratio of the first minor eigenvalue to the dominant eigenvalue (63), where *q* ranges from −1, representing completely disassortative mixing, to 1, completely assortative mixing.

### Transmission models

To explore the expected transmission dynamics under different assumptions of social mixing patterns, we used an age-structured stochastic Susceptible-Exposed-Infectious-Recovered (SEIR) transmission model to age-specific attack rates (*Supplementary Methods*). We used transmission parameters consistent with influenza taken from the literature (64) and simulated multiple stochastic realizations of a single outbreak in a closed population. Recognizing that proximity sensors and self-reported surveys were likely to record contacts with different transmission potential, we fitted *β* for each set of parameters, including the age-specific mixing matrix, using the next-generation matrix to give an *R*_0_ of 2.0, a moderate estimate of pandemic influenza (64, 65) (*Supplementary Methods*). Stochastic simulations were conducted using Gillespie’s direct algorithm.

### Ethical considerations and approvals

Informed consent was obtained through an opt-out process, where parents and legal guardians of students in participating schools were sent study information (including an opt-out declaration to return), prior to study activities and data collection in the schools. All study design of The SMART study, including the opt-out consenting process, was approved by the ethics committees at the University of Pittsburgh (PRO1102050), the University of Florida (IRB201701941), the University of Liverpool, and the Centers for Disease Control and Prevention (IRB00000319). Participating students were able to opt out at any time by simply saying that they did not wish to participate in a study activity. Non-participating students were given the option to wear non-operative electronic sensors to avoid any stigma associated with not participating in study activity. All research was performed in accordance with the protocol approved by the above institutions and in accordance with the relevant guidelines and regulations.

### Data availability

De-identified dataset is available through the Zenodo Repository doi:10.5281/zenodo.3940772.

## Data Availability

De-identified dataset is available through the Zenodo Repository doi:10.5281/zenodo.3940772

https://zenodo.org/record/3940772

## Acknowledgements

We are sincerely grateful to all students, teachers, administrators, and school district officials for their participation in and support of the SMART study. This research was supported by the US Centers for Disease Control and Prevention (CDC Cooperative Agreement 1U01CK00179-01). DATC and KHG received additional support from the US NIH MIDAS program U54 GM088491. JMR acknowledges support from the Engineering and Physical Sciences Research Council (EP/N014499/1). The findings and conclusions in this report are those of the authors and do not necessarily represent the official position of CDC.

## Additional Information

We declare no competing interests.

## Author Contributions

KHG, DATC, SZ, CVJ, JMR designed research. DATC, SZ, CVJ, DG, MLS, HG, JC, CL, GMHY, KL, PC, EN, JMR performed research. KHG, DATC, JMR analysed data and wrote manuscript. All authors reviewed the manuscript.

## Figures and Tables

Appendix 1. Contact survey administered to select participants in elementary schools (grades K to 5), Pittsburgh PA, USA, 2012

Appendix 2. Contact survey given to participants in middle and high school (grades 6 to 12) and select participants in elementary schools (grades K to 5), Pittsburgh PA, USA, 2012

